# Industry and workplace characteristics associated with the use of a COVID-19 contact tracing app in Japan: a nation-wide employee survey

**DOI:** 10.1101/2021.04.01.21254744

**Authors:** Tomohiro Ishimaru, Koki Ibayashi, Masako Nagata, Ayako Hino, Seiichiro Tateishi, Mayumi Tsuji, Akira Ogami, Shinya Matsuda, Yoshihisa Fujino, for the CORoNaWork Project

**Affiliations:** Department of Environmental Epidemiology, Institute of Industrial Ecological Sciences, University of Occupational and Environmental Health, Japan, Kitakyushu, Japan; Department of Occupational Health Practice and Management, Institute of Industrial Ecological Sciences, University of Occupational and Environmental Health, Japan, Kitakyushu, Japan; Department of Mental Health, Institute of Industrial Ecological Sciences, University of Occupational and Environmental Health, Japan, Kitakyushu, Japan; Department of Occupational Medicine, School of Medicine, University of Occupational and Environmental Health, Japan, Kitakyushu, Japan; Department of Environmental Health, School of Medicine, University of Occupational and Environmental Health, Japan, Kitakyushu, Japan; Department of Work Systems and Health, Institute of Industrial Ecological Sciences, University of Occupational and Environmental Health, Japan, Kitakyushu, Japan; Department of Preventive Medicine and Community Health, School of Medicine, University of Occupational and Environmental Health, Japan, Kitakyushu, Japan

**Keywords:** Contact tracing, COVID-19, SARS-CoV-2, smartphone, worksite

## Abstract

**Objectives:** To combat coronavirus disease 2019 (COVID-19), many countries have used contact tracing apps, including Japan’s voluntary-use contact-confirming application (COCOA). The current study aimed to identify industry and workplace characteristics associated with the use of this COVID-19 contact tracing app.

**Methods:** This cross-sectional study of full-time workers used an online survey. Multiple logistic regression analysis was used to evaluate the associations of industry and workplace characteristics with contact tracing app use.

**Results:** Of the 27,036 participants, 25.1% had downloaded the COCOA. Workers in the public service (adjusted odds ratio [aOR] = 1.29, 95% confidence interval [CI]: 1.14–1.45) and information technology (aOR = 1.38, 95% CI: 1.20–1.58) industries were more likely to use the app than were those in the manufacturing industry. In contrast, app usage was less common among workers in the retail and wholesale (aOR = 0.87, 95% CI: 0.76–0.99) and food/beverage (aOR = 0.81, 95% CI: 0.70–0.94) industries, but further adjustment for business size attenuated these associations. Workers at larger companies were more likely to use the app. Compared with permanent employees, the odds of using the app were higher for managers and civil servants but lower for those who were self-employed.

**Conclusions:** One possible reason for the under-implementation of the contact tracing app in the retail and wholesale and food/beverage industries is small business size, as suggested by the fully adjusted model results. An awareness campaign should be conducted to promote the widespread use of the contact tracing app in these industries.

## Introduction

A number of countries have used digital contact tracing tools to combat coronavirus disease 2019 (COVID-19) by tracing possible infected cases to prevent the spread of the disease.^1, 2^ Japan’s government released a contact tracing app for smartphones—the contact-confirming application (COCOA)—on June 19, 2020.^3^ Downloading the COCOA is not mandatory but it is recommended by authorities. As of March 18, 2021, the COCOA had been downloaded by 20.8% of Japanese citizens (26 million downloads) and used for 2.5% of positive cases (11,513/452,863).^3^ The COCOA uses Bluetooth to collect information on close contacts—those who have been within 1 meter of an individual who has installed the app for a duration of at least 15 minutes in the last 14 days. When a user is infected with COVID-19, they can alert others with whom they have been in close contact through the app. No personal information is stored in the central database. The notified person can then get tested at the nearest COVID-19 testing center.

High utilization is important for contact tracing apps because they are only effective when both those who are infected with COVID-19 and their close contacts have installed the app. According to a simulation study, 90% of the population needs to use the COCOA for effective infection control.^4^ In other countries, contact tracing apps, such as TraceTogether in Singapore, have frequently been used with mandatory installation and use.^2^ In contrast, this practice is less frequent in countries where smartphones are not widely used.^2^ In terms of using contact tracing apps, migrant workers, persons with low income, and older adults who may find the technology necessary to use the app difficult are all at a disadvantage.^5^ The acceptance rate of contact tracing apps has been found to range from 42% to 70% in developed countries.^6-8^ Several factors influence this acceptance: trust in the government, health concerns, privacy, battery usage, and concerns about the effectiveness of the app.^7^

In Japan, employers have promoted the use the COCOA among workers for effective contact tracing to prevent workplace COVID-19 clusters.^9^ Many workers are at risk of COVID-19 infection because of their close contact with colleagues or customers. However, there is little evidence about the associations of industry and workplace characteristics with the use of contact tracing apps. Promoting the use of contact tracing apps in the workplace is also important for preventing the spread of clusters from the working-age population to high-risk populations, such as older adults. To develop a strategy for the widespread use of contact tracing apps, it is necessary to identify the industries or workplaces in which such apps are under-implemented. The purpose of the current study was to identify which industry and workplace characteristics were associated with the use of a COVID-19 contact tracing app.

## Subjects and Methods

### Study design and participants

The current study was conducted as part of the Collaborative Online Research on the Novel-coronavirus and Work (CORoNaWork) Project. Details of the study protocol have been published elsewhere.^10^ Briefly, the CORoNaWork Project was an online cohort study in Japan. We extracted online self-administered questionnaire data from the baseline dataset, collected December 22–26, 2020, for cross-sectional analysis. Panelists who had registered with an online research company and who were currently working full-time were invited to participate in the survey. Health care workers and caregivers were not invited to participate. We selected 33,087 participants through cluster sampling stratified by sex, region, and job type. We excluded invalid responses, leaving data on 27,036 participants for analysis. When the baseline survey was conducted, the numbers of COVID-19 infections and deaths were much higher than they had been during the first and second waves of the disease in Japan; therefore, Japan was on maximum alert during this third wave. This study was approved by the Ethics Committee of the University of Occupational and Environmental Health, Japan (R2-079).

### Questionnaire

This study used questionnaire data including variables on (a) demographic characteristics; (b) health behavior; (c) risk perception; (d) industry and workplace characteristics; and (e) the outcome (use of the contact tracing app). The demographic variables included sex, age, marital status, education, and annual household income. The health behavior variables were smoking and regular alcohol intake. Risk perception was assessed using a single question on anxiety about contracting COVID-19, with the response options of *yes* and *no*. Three variables were used to assess industry and workplace characteristics: type of industry, business size, and occupation. Type of industry followed the Japan Standard Industrial Classification, with industries accounting for less than 3% of the sample categorized as “other.”^11^ Business size was classified as < 10, 10–49, 50–999, or ≥ 1,000 employees. Each participant reported their occupation as *permanent employee, manager, civil servant, dispatched or contract worker, self-employed*, or *other*. The outcome variable, the use of the contact tracing app, was assessed by asking “Have you downloaded the COCOA?” The response options were *yes* and *no*.

### Data analysis

Univariate and multiple logistic regression analyses were used to evaluate the associations of occupational factors with the use of the contact tracing app. We assessed three occupational factors as explanatory variables: type of industry, business size, and occupation. Three models were evaluated. Model 1 was the univariate model. Model 2 was adjusted for demographic, health behavior, and risk perception variables (sex, age, marital status, education, annual household income, smoking, alcohol intake, and anxiety about contracting COVID-19). Model 3 additionally adjusted for type of industry and business size. We did not include the occupation variables in Model 3 because of the high correlations between the occupation of civil servant and the public service industry and between self-employed occupation and business size of 1–9 employees. All *P*-values were two-sided, and statistical significance was set at *P* < 0.05. We used Stata/SE 16.1 (StataCorp, College Station, TX, USA) for all analyses.

## Results

Table 1 shows the general characteristics of the study participants. Of the 27,036 participants, 6,786 (25.1%) reported having downloaded the COCOA. Approximately half of the participants were women (48.9%), and about half were married (55.6%). The most frequently observed category for annual household income was 4–8 million JPY (44.1%; USD 1 = JPY 106.78 as of 2020).^12^ Excluding the “other” industry category, manufacturing accounted for the largest percentage of the sample (17.0%), followed by medical and welfare (16.6%) and public service (7.0%). Establishments with 50–999 employees (35.9%) and permanent employee (46.5%) were the most frequently observed business size and occupation categories, respectively.

**Table 1.** General characteristics of the study participants

|  | Total<br>*N* = 27,036<br>*n* (%) | COVID-19 contact tracing app users<br>*n* = 6,786 (25.1%) |  |
| --- | --- | --- | --- |
|  |  | *n* (%) | Utilization rate (%) |
| Sex |  |  |  |
| Female | 13,222 (48.9) | 3,181 (46.9) | 24.1 |
| Male | 13,814 (51.1) | 3,605 (53.1) | 26.1 |
| Age |  |  |  |
| 20–39 years | 6,763 (25.0) | 1,669 (24.6) | 24.7 |
| 40–49 years | 8,011 (29.6) | 1,886 (27.8) | 23.5 |
| 50–59 years | 9,012 (33.4) | 2,359 (34.8) | 26.2 |
| 60–65 years | 3,250 (12.0) | 872 (12.8) | 26.8 |
| Marital status |  |  |  |
| Single | 9,164 (33.9) | 1,991 (29.4) | 21.7 |
| Divorced or widowed | 2,843 (10.5) | 722 (10.6) | 25.4 |
| Married | 15,029 (55.6) | 4,073 (60.0) | 27.1 |
| Education |  |  |  |
| Junior high or high school | 7,321 (27.1) | 1,647 (24.3) | 22.5 |
| Vocational school or college | 6,544 (24.2) | 1,552 (22.9) | 23.7 |
| University or graduate school | 13,171 (48.7) | 3,587 (52.8) | 27.2 |
| Annual household income |  |  |  |
| < 2,000,000 Japanese yen | 1,709 (6.3) | 277 (4.1) | 16.2 |
| 2,000,000–3,999,999 Japanese yen | 5,548 (20.5) | 1,188 (17.5) | 21.4 |
| 4,000,000–7,999,999 Japanese yen | 11,927 (44.1) | 2,925 (43.1) | 24.5 |
| ≥ 8,000,000 Japanese yen | 7,852 (29.1) | 2,396 (35.3) | 30.5 |
| Current smoking | 7,004 (25.9) | 1,740 (25.6) | 24.8 |
| Regular alcohol intake | 11,017 (40.8) | 2,901 (42.7) | 26.3 |
| Anxiety about contracting COVID-19 (yes) | 20,017 (74.0) | 5,737 (84.5) | 28.7 |
| Industry |  |  |  |
| Manufacturing | 4,591 (17.0) | 1,165 (17.2) | 25.4 |
| Public service | 1,891 (7.0) | 614 (9.0) | 32.5 |
| Information technology | 1,331 (4.9) | 429 (6.3) | 32.2 |
| Retail and wholesale | 1,797 (6.6) | 394 (5.8) | 21.9 |
| Food/beverage | 1,319 (4.9) | 269 (4.0) | 20.4 |
| Medical and welfare | 4,500 (16.6) | 1,071 (15.8) | 23.8 |
| Finance | 1,226 (4.5) | 364 (5.4) | 29.7 |
| Construction | 944 (3.5) | 240 (3.5) | 25.4 |
| Other | 9,437 (34.9) | 2,240 (33.0) | 23.7 |
| Business size |  |  |  |
| 1–9 employees | 6,165 (22.8) | 1,272 (18.7) | 20.6 |
| 10–49 employees | 4,390 (16.2) | 957 (14.1) | 21.8 |
| 50–999 employees | 9,703 (35.9) | 2,491 (36.7) | 25.7 |
| ≥ 1,000 employees | 6,778 (25.1) | 2,066 (30.4) | 30.5 |
| Occupation |  |  |  |
| Permanent employee | 12,575 (46.5) | 3,006 (44.3) | 23.9 |
| Manager | 3,403 (12.6) | 1,160 (17.1) | 34.1 |
| Civil servant | 2,810 (10.4) | 840 (12.4) | 29.9 |
| Dispatched or contract worker | 2,894 (10.7) | 671 (9.9) | 23.2 |
| Self-employed | 2,819 (10.4) | 525 (7.7) | 18.6 |
| Other | 2,535 (9.4) | 584 (8.6) | 23.0 |

Table 2 shows the associations of industry and workplace characteristics with the use of the COVID-19 contact tracing app. Participants in the public service (adjusted odds ratio [aOR] = 1.29, 95% *c*onfidence interval [CI]: 1.14–1.45, Model 3) and information technology (aOR = 1.38, 95% CI: 1.20–1.58, Model 3) industries were more likely to use the app than were those in the manufacturing industry. In contrast, app usage was less common among participants working in the retail and wholesale and food/beverage industries, and adjusting for demographic, health behavior, and risk perception variables did not alter these associations remarkably (retail and wholesale: aOR = 0.87, 95% CI: 0.76–0.99; food/beverage: aOR = 0.81, 95% CI: 0.70–0.94; Model 2); however, after further adjustment for business size in Model 3, these industry types showed no significant difference from the manufacturing industry.

**Table 2.** Associations between occupational factors and the use of the COVID-19 contact tracing app

|  | Model 1 |  |  | Model 2 |  |  | Model 3 |  |  |
| --- | --- | --- | --- | --- | --- | --- | --- | --- | --- |
|  | OR | (95% CI) | p value | OR | (95% CI) | P-value | OR | (95% CI) | P-value |
| Industry |  |  |  |  |  |  |  |  |  |
| Manufacturing | 1.00 | - | - | 1.00 | - | - | 1.00 | - | - |
| Public service | 1.41 | (1.25–1.58) | < 0.001 | 1.32 | (1.17–1.49) | < 0.001 | 1.29 | (1.14–1.45) | < 0.001 |
| Information technology | 1.39 | (1.21–1.58) | < 0.001 | 1.36 | (1.18–1.55) | < 0.001 | 1.38 | (1.20–1.58) | < 0.001 |
| Retail and wholesale | 0.83 | (0.73–0.94) | 0.004 | 0.87 | (0.76–0.99) | 0.034 | 0.92 | (0.81–1.05) | 0.223 |
| Food/beverage | 0.76 | (0.66–0.89) | < 0.001 | 0.81 | (0.70–0.94) | 0.007 | 0.89 | (0.76–1.03) | 0.125 |
| Medical and welfare | 0.94 | (0.85–1.03) | 0.201 | 0.86 | (0.78–0.94) | 0.004 | 0.91 | (0.82–1.01) | 0.068 |
| Finance | 1.26 | (1.09–1.45) | 0.001 | 1.19 | (1.03–1.38) | 0.016 | 1.15 | (0.99–1.32) | 0.061 |
| Construction | 1.00 | (0.85–1.18) | 0.999 | 0.98 | (0.84–1.16) | 0.845 | 1.10 | (0.93–1.30) | 0.271 |
| Other | 0.92 | (0.84–0.99) | 0.034 | 0.94 | (0.87–1.03) | 0.175 | 1.01 | (0.93–1.10) | 0.807 |
| Business size |  |  |  |  |  |  |  |  |  |
| 1–9 employees | 1.00 | - | - | 1.00 | - | - | 1.00 | - | - |
| 10–49 employees | 1.09 | (0.99–1.20) | 0.063 | 1.04 | (0.94–1.14) | 0.453 | 1.03 | (0.97–1.11) | 0.329 |
| 50–999 employees | 1.36 | (1.25–1.46) | < 0.001 | 1.24 | (1.14–1.34) | < 0.001 | 1.22 | (1.13–1.33) | < 0.001 |
| ≥ 1,000 employees | 1.71 | (1.57–1.85) | < 0.001 | 1.52 | (1.39–1.65) | < 0.001 | 1.44 | (1.32–1.57) | < 0.001 |
| Occupation* |  |  |  |  |  |  |  |  |  |
| Permanent employee | 1.00 | - | - | 1.00 | - | - |  |  |  |
| Manager | 1.62 | (1.48–1.76) | < 0.001 | 1.40 | (1.28–1.53) | < 0.001 |  |  |  |
| Civil servant | 1.34 | (1.23–1.47) | < 0.001 | 1.22 | (1.11–1.34) | < 0.001 |  |  |  |
| Dispatched or contact worker | 0.95 | (0.86–1.04) | 0.277 | 1.02 | (0.92–1.12) | 0.769 |  |  |  |
| Self-employed | 0.72 | (0.64–0.80) | < 0.001 | 0.76 | (0.69–0.85) | < 0.001 |  |  |  |
| Other | 0.95 | (0.86–1.05) | 0.290 | 0.89 | (0.80–0.99) | 0.033 |  |  |  |

COVID-19: coronavirus disease 2019; OR: odds ratio; CI: confidence interval
Model 1: Adjusted for sex and age
Model 2: Adjusted for sex, age, income, marital status, education, smoking, alcohol intake, and anxiety about contracting COVID-19
Model 3: Adjusted for sex, age, income, marital status, education, smoking, alcohol intake, anxiety about contracting COVID-19, type of industry, and business size
\* The occupation variables were not included in Model 3 because of the high correlations between the occupation of civil servant and the public service industry and between self-employed occupation and business size of 1–9 employees.

Participants in larger companies were more likely to use the app (50–999 employees: aOR = 1.21, 95% CI: 1.12–1.32; ≥ 1,000 employees: aOR = 1.37, 95% CI: 1.24–1.52; Model 3). Regarding occupation, compared with permanent employees, the odds of app usage were higher for managers (aOR = 1.40, 95% CI: 1.28–1.53, Model 2) and civil servants (aOR = 1.22, 95% CI: 1.11–1.34, Model 2) but lower for those who were self-employed (aOR = 0.76; 95% CI: 0.69–0.85, Model 2).

## Discussion

The current study identified that working in the public service sector, in the information technology industry, or for a business with 50 or more employees, as well as holding the position of manager or civil servant, were factors facilitating the use of the contact tracing app—over 30% of each of these groups had downloaded the app. In contrast, working in the retail and wholesale and food/beverage industries and being self-employed were associated with a lower likelihood of using the app—around 20% of these groups had downloaded the app. These findings give insight needed for the development of a strategy for the widespread use of COVID-19 contact tracing apps.

An important finding of this study was that the contact tracing app was under-implemented in the retail and wholesale and food/beverage industries. This finding did not change substantially after adjusting for demographic, health behavior, and risk perception variables. However, further adjustment for business size attenuated these associations. One possible reason for this change in results is that most workplaces in these types of industries are small-scale, which may be the cause of insufficient use of the app. This idea is consistent with a previous study showing that small companies were relatively unlikely to implement workplace measures against COVID-19.^13^ This may be because of the limited opportunities to obtain information and the lack of human, time, and financial resources to take countermeasures against health threats at small companies.^14^ However, workers in these industries have particularly a strong need to use the contact tracing app because they are in contact with an unspecified number of people, such as in restaurants and bars, where customers eat and drink without masks.^15^ An awareness campaign should be conducted for the retail and wholesale and food/beverage industries to promote the use of the contact tracing app.^16^

We found that the contact tracing app was frequently used by civil servants and managers. This finding implies that civil servants and managers might follow requests to use the app made through organized channels because workers were encouraged to use the COCOA by the government and by employer associations.^9^ A previous study conducted in Japan reported that the public service sector most frequently received announcements regarding measures to prevent COVID-19 in the early stage of the pandemic^17^—this also suggests that the public service sector provides COVID-19 advisory information to civil servants. These results indicate that app uptake may increase when workers, including managers, receive messages through organized channels, even if the requests are voluntary. For non-managers, a sharing function allowing users to invite coworkers and friends to download the app may increase the rate of installation of the app.^7^

Workers in the information technology industry, such as system engineers, have the advantage of expertise in digital technology and know how to use smartphone apps. The current study revealed that information technology industry workers used the contact tracing app more frequently, compared with workers in other industries. This finding is in line with a previous study showing that high information technology literacy eliminated technical obstacles to using contact tracing apps.^18^ In addition, the information technology industry actively implements workplace measures against COVID-19, such as teleworking, and these practices may also relate to the finding.^17^ These results suggest that a technical support service for those who have difficulty using the contact tracing app may improve app uptake.

This study has several limitations. First, the study recruited panelists who registered with an online research company. Therefore, the participants may not represent general workers. For example, online panelists may be particularly willing to use online tools or more familiar than others with using apps. Consequently, the results may underestimate negative factors for use of the app. Second, we evaluated the use of the contact tracing app by asking participants about whether they had downloaded it. Therefore, we did not confirm whether the application was installed. However, we think that most people who downloaded the app also installed it. Despite these limitations, to the best of our knowledge, this study represents the first study in Japan to examine current use of the COCOA with a large sample.

In conclusion, the present study evaluated the associations of industry and workplace characteristics with the use of a COVID-19 contact tracing app in a large-scale online survey of Japanese workers. Those working in the public service sector or in information technology, as well as managers, were frequently found to use the contact tracing app, whereas those working in the retail and wholesale and food/beverage industries were less likely to use it. One possible reason for the under-implementation of the contact tracing app in the retail and wholesale and food/beverage industries may be the small size of businesses in these types of industries. An awareness campaign should be conducted for workers in these industries to promote the widespread use of the contact tracing app to help these workers trace possible infected cases, preventing the spread of the disease.

## Data Availability

No additional data are available.

## Acknowledgments

This study was supported by a research fund from the University of Occupational and Environmental Health, Japan; General Incorporated Foundation (Anshin Zaidan): The Development of Educational Materials on Mental Health Measures for Managers at Small-sized Enterprises; Health, Labour and Welfare Sciences Research Grants: Comprehensive Research for Women’s Healthcare (H30-josei-ippan-002); Research for the Establishment of an Occupational Health System in Times of Disaster (H30-roudou-ippan-007), and scholarship donations from Chugai Pharmaceutical Co., Ltd.

The current members of the CORoNaWork Project, in alphabetical order, are as follows: Dr. Yoshihisa Fujino (present chairperson of the study group), Dr. Akira Ogami, Dr. Arisa Harada, Dr. Ayako Hino, Dr. Chimed-Ochir Odgerel, Dr. Hajime Ando, Dr. Hisashi Eguchi, Dr. Kazunori Ikegami, Dr. Keiji Muramatsu, Dr. Koji Mori, Dr. Kyoko Kitagawa, Dr. Masako Nagata, Dr. Mayumi Tsuji, Dr. Rie Tanaka, Dr. Ryutaro Matsugaki, Dr. Seiishiro Tateishi, Dr. Shinya Matsuda, Dr. Tomohiro Ishimaru, Dr. Tomohisa Nagata, Dr. Yosuke Mafune, and Ms. Ning Liu. All members are affiliated with the University of Occupational and Environmental Health, Japan.

## Disclosure

*Approval of the research protocol*: This study was approved by the Ethics Committee of the University of Occupational and Environmental Health, Japan (R2-079). *Informed Consent*: Informed consent was obtained from all participants. *Registry and the Registration No. of the study/trial*: N/A. *Animal Studies*: N/A. *Conflict of Interest*: N/A.

## Notes

### Competing Interest Statement

The authors have declared no competing interest.

### Author Declarations

This study was approved by the Ethics Committee of the University of Occupational and Environmental Health, Japan (R2-079).

